# Joint Impacts of Nutrition and Socioeconomic Status on SARS-CoV-2 Antibody Titer in a Prospective Birth Cohort in Brazil

**DOI:** 10.1101/2024.08.22.24312306

**Authors:** Moin Vahora, Otávio Leão, Mariângela Freitas da Silveira, Marlos Rodrigues Domingues, Pedro Hallal, Alicia N.M. Kraay

## Abstract

**Purpose:** In this study, we examine the joint effects of stunting and socioeconomic status (SES) on COVID-19 serostatus in Brazil and whether these patterns vary by COVID-19 vaccine status.

**Methods:** We use data from birth through the 7-year follow-up of the Pelotas 2015 birth cohort, including anthropometry, demographics, vaccination status, and COVID-19 antibody test results. We use linear regression models to examine the associations between SES (exposure) and stunting (effect modifier) on COVID-19 antibody titer among children who had not been vaccinated for COVID-19 (n=1,103) and among children who had been vaccinated (n=1,875).

**Results:** All SES strata had a statistically identical probability of testing positive. While not statistically significant, stunted children who were unvaccinated tended to have lower COVID-19 titers (difference -0.10, 95% CI: -0.21,0.004) compared with children who were not stunted, and this difference was attenuated among stunted children who were vaccinated. Duration of school enrollment was associated with increased antibody titer, with each month being associated with a 0.17 unit higher increase in OD titer (95% CI: 0.15, 0.18).

**Conclusions:** This result may suggest that stunted children have poorer immune responses to natural infection and thus lower immunity, but vaccination can overcome this deficit. Future recommendations include population-wide follow-up vaccination, particularly for stunted children. Given increasing antibody titer with a longer duration of school attendance, primary COVID-19 vaccination and potential boosters may be useful for children prior to school entry to reduce the risk of natural infection.

## Introduction

The global impact of COVID-19, an infectious viral respiratory disease, was significantly high, with Brazil being one of the nations with the highest incidence [1–3]. Brazil ranked third in total number of COVID-19 cases and second highest number of deaths [4]. COVID-19 was the number one cause of death in Brazil in March of 2021 according to the Brazil health metrics and evaluation briefing [5] and COVID remained a persistent problem in the country through June 2024 [1].

Several factors are known to increase the risk of severe COVID-19, with socioeconomic status (SES) being a major contributor. People with low SES are more likely to have increased exposure to COVID-19 due to their reduced capacity to social distance and are more likely to have severe outcomes following exposure because of limited healthcare and high stress associated with poverty [6]. For instance, increased stress is known to weaken the immune system and compromise immune health, leading to higher symptomatic infection rates [7–12]. Furthermore, economically disadvantaged people living in municipalities with overcrowded conditions are shown to have an elevated risk of mortality from COVID-19 [13–17]. Specifically, a study conducted in northeast Brazil found that people from low SES populations had higher mortality related to COVID-19 [18]. People with low SES are less likely to be fully vaccinated and have higher rates of COVID-19 infection. A study found that a higher coverage of two vaccine doses was achieved among children with high SES, compared to lower coverage among low SES households [19]. Even if they are ultimately vaccinated, individuals with low socioeconomic status are also often delayed in receiving their vaccine doses in the initial phases of vaccine rollout and may be less likely to complete the series [20].

Poor nutritional status from low SES has previously been linked to children being immunocompromised and subsequently having a higher severity of COVID-19. Children with lower SES experience a higher prevalence of poor anthropometric status, including stunting and wasting, than children with higher SES [21]. In addition, the highest prevalence of stunting and wasting is seen in low-and middle-income countries (LMIC) [22]. The population of people with low SES status increased during the pandemic due to lost jobs and people have alternatively utilized less expensive sources of calories. These alternative food sources are less nutrient-rich, resulting in an increasing prevalence of malnutrition [23–25]. Malnutrition from micronutrient deficiencies is a significant risk factor for fatal COVID-19 infection in children, especially in low- and middle-income countries [26]. In contrast, high nutritional quality was found to be associated with a lower risk of COVID-19 severity [27]. More importantly, stunting and other indicators of poor nutritional status have been associated with more frequent infections. For example, stunted children are two to four times more likely to die before the age of five than their counterparts due to an impaired immune function being at greater risk of infectious mortality and morbidity [28–30]. Children who are stunted are ultimately more likely to suffer from infectious diseases and form new infections than non-stunted children.

Although COVID-19 infections have impacted all populations, much less is known about the impact of COVID-19 in children than adults. Current studies have focused on the impact of severe diseases affecting children, but there is limited knowledge about infection rates and immunity [31]. For example, a study focusing on health outcomes in children found that children had milder symptoms than adults, but a subset of children developed severe diseases [32].

Additionally, there is limited data available on the incidence and potential risk factors associated with infection. Diverse sources suggest there is much unknown about the rate of COVID-19 in children due to low testing and surveillance, in part because of their milder symptoms [32–34]. The presence and level of antibodies against COVID-19 provide one useful metric of COVID-19 immunity and burden, particularly in regions with low access to or low utilization of testing [35,36]. Therefore, in this study, we used IgG antibody positivity and titer to better assess recent infection burden.

The current study uses data from the seven-year follow-up of the 2015 Pelotas Birth Cohorts to assess the association between socioeconomic status (SES) and COVID-19 seropositivity in Pelotas, Brazil, and whether this effect is modified by nutritional status and/or prior vaccination [37]. In this study, OD titers and seropositivity are used as a proxy of current immunity and recent infection. The present study investigates the relationship between SES (primary exposure) and COVID-19 seropositivity (outcome), and how this association might be modified by nutrition and vaccination (effect modifiers).

### Method

### Study population

The data used for this study came from the Pelotas (Brazil) birth cohort study, which provides information on time trends in areas such as maternal and child health, nutrition, socioeconomic status, physical activity, and related factors [37]. Pelotas is a diverse, mid-sized city in the south of Brazil [37]. All children delivered in the city hospitals of 1982, 1993, 2004, and 2015 were enrolled at birth, with frequent follow-up visits during childhood and with less frequent visits thereafter. For this specific analysis, we use data from the 2015 cohort from birth until the 7-year follow-up (in 2022). At each study visit, the mothers were interviewed based on a standardized computer-assisted questionnaire approved by the ethics committee at the Federal University of Pelotas. In the 7-year assessment, the data was collected through interviews, biological samples, and anthropometric measurements. In our current analysis, we utilized interview questions related to socioeconomic status such as “*What was the income of each family member?*”, and anthropometry measures (weight/height).

### Variables

#### COVID-19 outcomes

We considered two COVID-19 outcomes in our analysis: seropositivity (yes/no), and antibody titer (in OD units). We used data on current antibody titers to assess past infection with COVID-19. The current antibody titer was assessed using a blood spot assay validated for the current cohort to measure IgG titer, measured in optical density [38]. Similar assays have been used in other populations to assess population immunity [31]. IgG antibodies wane relatively rapidly following natural infection, so they are most useful for capturing recent infection [38]. Antibodies may persist for longer following severe infection, so antibody positivity provides data about both past infection severity and its recency. OD titer data was subsequently categorized into positive/negative values based on established criteria [37–39]. In this analysis, we use both the continuous version (OD titer) and binary (positive/negative). We focus on the OD titer in the main text (linear regression) but include analysis with the binary version of seroprevalence (logistic regression) in the supplementary materials.

### Exposures

In this analysis, we focused on three exposures of interest (SES, undernutrition, and COVID-19 vaccine status). Undernutrition was the primary effect modifier, with SES and COVID-19 status being hypothesized to modify the association between undernutrition and seropositivity.

#### Undernutrition (Exposure)

We used information on child weight and length at study visits to calculate length for age z-scores, and weight for length/height z-scores for each follow-up. Children who were more than two standard deviations below the weight for height z-score at *any* follow-up visit were classified as wasted (n=128) and children who were more than two standard deviations below the length for age z-score at *any* follow-up visit were classified as stunted (n=410) per WHO definitions [40]. Because we did not have sufficient power to analyze wasted children separately and because stunting and wasting were collinear, we focused on stunting as our primary nutrition indicator for this analysis.

#### Socioeconomic (SES) Status (Effect Modifier)

The total household income in *Reais* (Brazilian currency) at the 2021/2022 seven-year follow-up visit was used to define SES. Total family income was divided into quintiles with the lowest quintile being used as the reference.

#### COVID-19 Vaccination (Effect modifier a priori)

At the 7-year follow-up visit, parents were asked whether the child was vaccinated against COVID-19. Because vaccination causes IgG antibodies to be developed that would be detected by the OD assay but are not indicative of natural infection, children who were vaccinated against COVID-19 (n=1,853) were analyzed separately from unvaccinated children (n=1,103) for all regression analyses, which enabled us to determine if the associations between SES/stunting and antibody titer/positivity varied by vaccine status.

### Covariates

Parents of child cohort members were asked if their child had received the triple bacterial vaccine (DTP) and how many doses. While full vaccination is 6 or more doses (the standard series), we considered the number of doses received as the primary covariate. The level of DTP vaccination was chosen as a proxy for access to routine healthcare among cohort members, which might be a common cause of COVID-19 diagnosis and child nutritional status. We also included age at the seven-year follow-up, gender (male/female), and months since school entry. All children began school on March 1, 2022, so the months since entry were based on the number of months that had elapsed between school entry and the follow-up visit date.

## Statistical Analysis

### Descriptive Statistics

First, we performed descriptive analysis on socioeconomic status, stunting status, COVID-19 seropositivity, mothers’ education, DTP vaccine status, gender, months since school entry, and child age for this study.

### Linear regression model description

In our study, the primary outcome variable was OD titer for COVID-19 as it increased our statistical power and examined potentially important trends in antibody titer. The study’s predictors included socioeconomic status, stunting, age, school entry (months), and immunization status (DTP). All variables were included in the fully adjusted regression model. We presented only the unadjusted and adjusted model as the interaction terms were not significant. While our main analysis is stratified by COVID-19 vaccine status, we also combined all children into one model and tested for interactions between COVID-19 vaccine status and stunting.

### Sensitivity analysis: Logistic regression model description

We also repeated the main linear regression models using COVID-19 positivity (yes/no) as the outcome of interest, running as a logistic regression. The outcome variable for logistic regression was whether the children tested positive or negative for COVID-19, which was classified as 1 = positive and 0 = negative COVID-19 result. The study’s predictors included socioeconomic status, stunting, age, school entry (months), and immunization status (DTP).

### Software

All statistical analysis was conducted using STATA version 17.

## Results

### Descriptive statistics

Characteristics of children in the study, stratified by COVID-19 vaccine receipt, are shown in Table 1. Overall, vaccinated (n=1,853) and unvaccinated (n=1,132) children were similar apart from COVID antibody titer. OD titer was higher among vaccinated children than unvaccinated children (OD for vaccinated=1.70, SD=0.69; OD for unvaccinated=1.22, SD=0.55). Seropositivity was also higher among the vaccinated children (87%, n=175) compared with unvaccinated children (42%, n=358). Vaccination receipt seemed to be approximately even across SES strata (Table 1). Vaccinated and unvaccinated children were also equally likely to be male. Furthermore, because of data sparsity and the low prevalence of malnutrition in our sample, we established ever-stunted and ever-wasted variables for stunting and wasting status, respectively. Overall, about 4% of participants had been ever wasted and 13% had been ever stunted, and this history of malnutrition was similar for vaccinated and unvaccinated children. For immunization, the number of DTP vaccine doses received was analyzed as a continuous variable based on the distribution of the variable (i.e., less than 1% of the participants received six or more doses). On average, vaccinated children received 2.81 doses of DTP and 2.68 for unvaccinated children. See the appendix for more details on the distribution of variables.

**Table 1.** Demographic characteristics of children in the 2015 cohort, comparing children who were vaccinated for COVID-19 with unvaccinated children. The categorical variables are shown as n (%), and continuous variables are shown as mean (SD). *Indicates a statistically significant difference between vaccinated and unvaccinated children. Titer and positivity results are shown in Figures S1 and S2.

|  | **Unvaccinated**<br>**(n=1,132)**<br>**N (%)** | **Vaccinated**<br>**(n=1,875)**<br>**N (%)** |
| --- | --- | --- |
| **Gender** |  |  |
| Male | 565 (49.91%) | 949 (50.61%) |
| Female | 567 (50.09%) | 926 (49.39%) |
| **SES (Family Income in quintiles)** |  |  |
| 1 (low) | 215 (19.16%) | 402 (21.73%) |
| 2 | 244 (21.75%) | 359 (19.41%) |
| 3 | 226 (20.14%) | 353 (19.08%) |
| 4 | 212 (18.89%) | 343 (18.54%) |
| 5 (high) | 225 (20.05%) | 393 (21.24%) |
| **Ever stunted** |  |  |
| No | 983 (86.84%) | 1,613 (86.07%) |
| Yes | 149 (13.16%) | 261 (13.93%) |
| **Ever wasted** |  |  |
| No | 1,084 (96.01%) | 1,786 (95.56%) |
| Yes | 45 (3.99%) | 83 (4.44%) |
| **Undernourished (stunted and/or wasted)** |  |  |
| No | 952 (84.32%) | 1544 (82.61%) |
| Yes | 177 (15.68%) | 325 (17.39%) |
| **COVID-19 Results** |  |  |
| Positive* | 487 (57.63%) | 1,220 (87.46%) |
| Negative | 358 (42.37%) | 175 (12.54%) |
| **Doses of DTP (triple bacterial vaccine)** | 2.68 (1.45) | 2.81 (1.49) |
| **Age (years)** | 6.79 (.32) | 6.84 (.34) |
| **OD titer** | 1.22 (.55) * | 1.70 (.69) * |
| **School Entry (months)** | 60.70 (76.06) | 87.27 (71.01) |

**Table 2.**
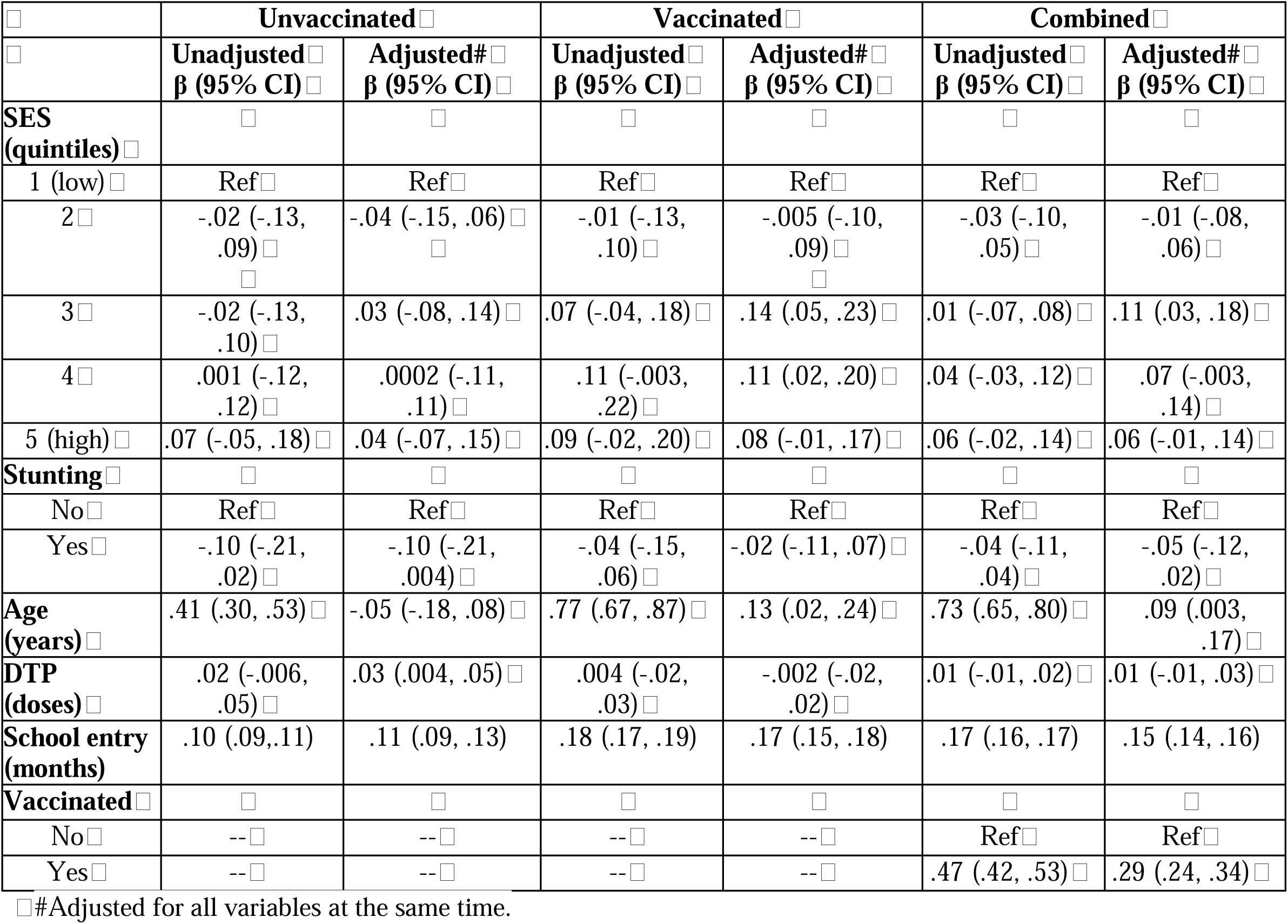
Linear regression showing an association between each variable and OD titer (outcome).

|  | Unvaccinated |  | Vaccinated |  | Combined |  |
| --- | --- | --- | --- | --- | --- | --- |
| | Unadjusted<br>$\beta$ (95% CI) | Adjusted#<br>$\beta$ (95% CI) | Unadjusted<br>$\beta$ (95% CI) | Adjusted#<br>$\beta$ (95% CI) | Unadjusted<br>$\beta$ (95% CI) | Adjusted#<br>$\beta$ (95% CI) |
| **SES (quintiles)** |  |  |  |  |  |  |
| 1 (low) | Ref | Ref | Ref | Ref | Ref | Ref |
| 2 | -.02 (-.13, .09) | -.04 (-.15, .06) | -.01 (-.13, .10) | -.005 (-.10, .09) | -.03 (-.10, .05) | -.01 (-.08, .06) |
| 3 | -.02 (-.13, .10) | .03 (-.08, .14) | .07 (-.04, .18) | .14 (.05, .23) | .01 (-.07, .08) | .11 (.03, .18) |
| 4 | .001 (-.12, .12) | .0002 (-.11, .11) | .11 (-.003, .22) | .11 (.02, .20) | .04 (-.03, .12) | .07 (-.003, .14) |
| 5 (high) | .07 (-.05, .18) | .04 (-.07, .15) | .09 (-.02, .20) | .08 (-.01, .17) | .06 (-.02, .14) | .06 (-.01, .14) |
| **Stunting** |  |  |  |  |  |  |
| No | Ref | Ref | Ref | Ref | Ref | Ref |
| Yes | -.10 (-.21, .02) | -.10 (-.21, .004) | -.04 (-.15, .06) | -.02 (-.11, .07) | -.04 (-.11, .04) | -.05 (-.12, .02) |
| **Age (years)** | .41 (.30, .53) | -.05 (-.18, .08) | .77 (.67, .87) | .13 (.02, .24) | .73 (.65, .80) | .09 (.003, .17) |
| **DTP (doses)** | .02 (-.006, .05) | .03 (.004, .05) | .004 (-.02, .03) | -.002 (-.02, .02) | .01 (-.01, .02) | .01 (-.01, .03) |
| **School entry (months)** | .10 (.09, .11) | .11 (.09, .13) | .18 (.17, .19) | .17 (.15, .18) | .17 (.16, .17) | .15 (.14, .16) |
| **Vaccinated** |  |  |  |  |  |  |
| No | -- | -- | -- | -- | Ref | Ref |
| Yes | -- | -- | -- | -- | .47 (.42, .53) | .29 (.24, .34) |

#Adjusted for all variables at the same time.

### Linear Regression

While not statistically significant, stunted children (primary exposure) tended to have lower antibody titers than children who had not previously been stunted, and this association was stronger among unvaccinated children. Specifically, unvaccinated children had an OD titer that was 0.10 units lower than children who had never been stunted (95% CI: -0.21, 0.004) whereas the difference was only 0.02 units among previously vaccinated children (95% CI: -0.11, .07). In the combined model, antibody titer was 0.29 units higher among vaccinated children than unvaccinated children (95% CI: 0.24, 0.34). SES was not associated with antibody titer. While age was positively associated with OD titer in the unadjusted model, this association disappeared after adjusting for the other variables, primarily due to the relationship with months since school entry. Each month increase in school attendance duration was associated with a significant rise in OD titer for both vaccinated (beta=0.17, 95% CI: 0.15, 0.18) and unvaccinated children (beta = 0.11, 95% CI: 0.09, 0.13). For unvaccinated children, number of DTP doses was positively associated with higher antibody titer, with each dose conferring a 0.03 unit increase in titer (95% CI: 0.004, 0.05), equivalent to a 0.18 unit increase for a child who had received the full DTP series compared with a child who had not received any doses. However, this association was not present among children who had been vaccinated for COVID-19 or in the combined model. Findings from the logistic regression were similar to the linear regression analysis (Table S1).

## Discussion

In summary, our findings suggest that stunting may impair immunity acquisition and/or durability among children, vaccination is associated with increased antibody titer among all children, regardless of their nutritional status, and may overcome any deficits. In contrast, SES had no association with COVID-19 seropositivity and/or titer. We also found similar distributions of SES for children with and without COVID-19 vaccination, implying that the uptake of vaccines in Brazil as of 2022 (during the omicron wave) was not different by SES status. Older children tended to have higher antibody titers and seropositivity than younger children, but this appeared to be largely related to school exposure. Specifically, children who had been in school for longer had increased antibody response compared to children not in school, potentially due to their increased exposure.

In the present study, we found that seroprevalence among unvaccinated children was independent of SES. We had initially expected that lower SES individuals would have a higher prevalence of infection due to increased exposure. However, it is estimated that more than 99% of the Brazilian population had been infected at the time of data collection [5]. As a result, in this highly exposed population, the pathway via exposure may be less important, and differences in seroprevalence are more likely related to the immune response to infection and the recency of exposure rather than cumulative risk.

While not significant, our findings related to undernutrition were our expectation based on other pathogens. Stunted children who were unvaccinated had lower antibody titers against COVID-19 compared with un-stunted children, with a similar but attenuated pattern among vaccinated children. The findings show the benefit of vaccination on higher titer even for stunted children, implying that vaccination may help overcome immune shortfalls for stunted children. The extent to which these benefits persist over time is unclear, particularly as new variants continue to emerge [41]. It is also important to note that although vaccination boosting might be beneficial, no children in our cohort had received booster vaccines, so we were not able to explore this pattern directly.

We also found that with increased age the children have higher odds of testing positive, even within the same age cohort (as all children were a similar age at testing) [2]. These findings could be explained by higher seropositivity with age, which could indicate improved seroconversion in older children. Alternatively, older children may have had higher prior exposure accrued prior to testing. Given that these children are approximately the same age, this is unlikely to be due to age-specific changes in behavior. However, this result is more likely to be influenced by whether children were in school as well as by calendar time.

We found strong evidence that the longer that the children were in school higher the antibody response (OD) titer. Similarly, logistic regression showed that the longer the children were in school, the higher their chance of testing positive. For example, vaccinated children had 60% increased odds of testing positive for COVID-19 per month of school exposure, and un-vaccinated children had 40% increased odds of testing positive. The linear regression model suggested that unvaccinated children with four months of school attendance had roughly similar titer to vaccinated children who had not yet begun attending school.

However, during late 2022 and early 2023, there was a wave of COVID-19 in Brazil that may have impacted all children even if they had not been in school [2,15]. Because all children in this cohort entered school at the same point in time, we cannot distinguish these two exposures and determine the separate effects of school vs. non-school exposure. In the United States, COVID-19 incidence has been shown to vary with the start and return to school following breaks, and thus these secular trends may also have been driven by school attendance [42–44]. In particular, the initial wave was generally coincident with the start of the school year in February, with a second smaller peak occurring around the typical winter break in July.

Our findings from this study demonstrate vaccine immunogenicity as vaccine receipt was associated with higher antibody titer and seropositivity and thus presumably immunity among children in Brazil. This result suggests that vaccination policies are likely effective in promoting immunity and promoting vaccination and ensuring equitable access are likely to provide population-level benefits. In addition, children who were in school longer had higher odds of testing positive for COVID-19, which may suggest that encouraging child vaccination in particular prior to school entry when exposure is likely to increase may be helpful.

One of the limitations of our study was the low prevalence of stunting in the sample, which decreased our statistical power. The trends illustrated here for titer and seropositivity might be statistically significant in a larger sample. Future studies could use other markers of undernutrition, with the goal of collecting data to understand the impact of COVID-19 infection on stunted and wasted children. We were also limited by the lack of data on the recency of exposure based on the date of last vaccination and/or infection and by the fact that seropositivity was only measured at a single time point, without specific surveillance data on natural infection. Future researchers could collect these data to better understand both the initial antibody response and time course of antibody waning and whether nutritional status might impact these trends. Such data might allow researchers to determine if vaccination can bring child immunity among stunted children up to the antibody titers seen among un-stunted children and might also be useful for assessing the optimal frequency of booster vaccination. Presently, booster doses for children are not recommended in Brazil and have also not been suggested by the World Health Organization [45]. Additional data on changes in antibody titers in children over time post-vaccination might be useful to see if adjustments to this policy are warranted, particularly prior to school entry.

Despite these weaknesses, our study has several strengths. Specifically, the prospective cohort study design followed the children since birth and included data on both nutritional status and COVID-specific exposures. The cohort study design allowed us to use data with follow-ups of a set of children selected in the 2015 cohort. Our study highlights the potential benefit of vaccination in a pediatric population in Brazil as a tool to both reduce overall incidence and narrow inequities based on prior nutritional status, particularly prior to school entry.

## Data Availability

All data produced in the present study are available upon reasonable request to the authors.

## Appendices

**Supplementary Table 1.**
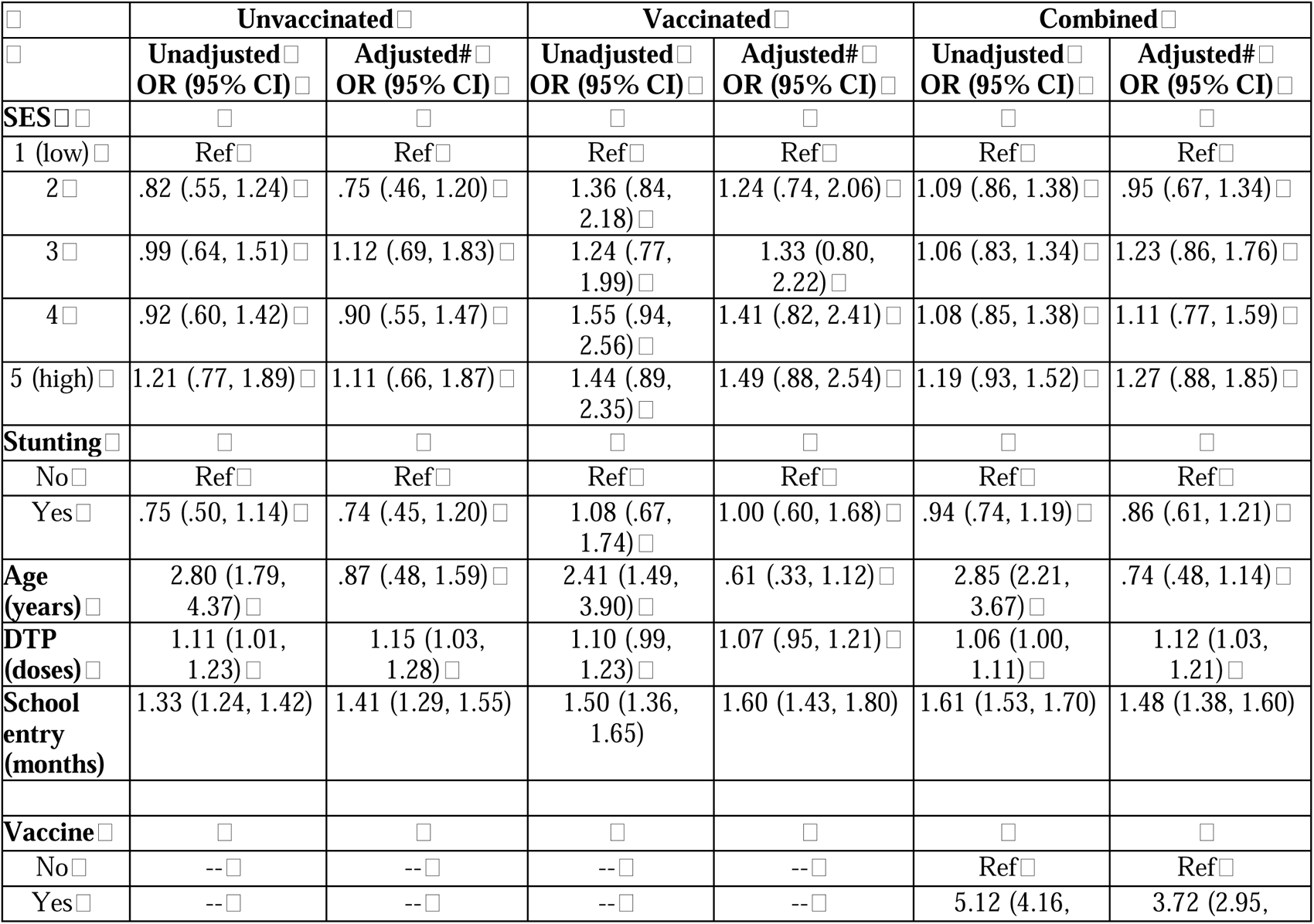
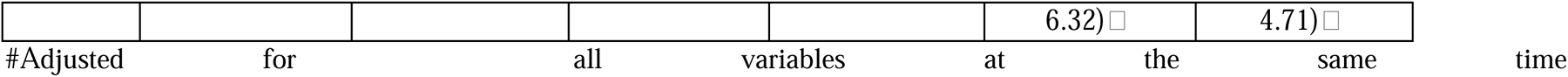
Logistic regression.

**Supplementary Figure 1:**
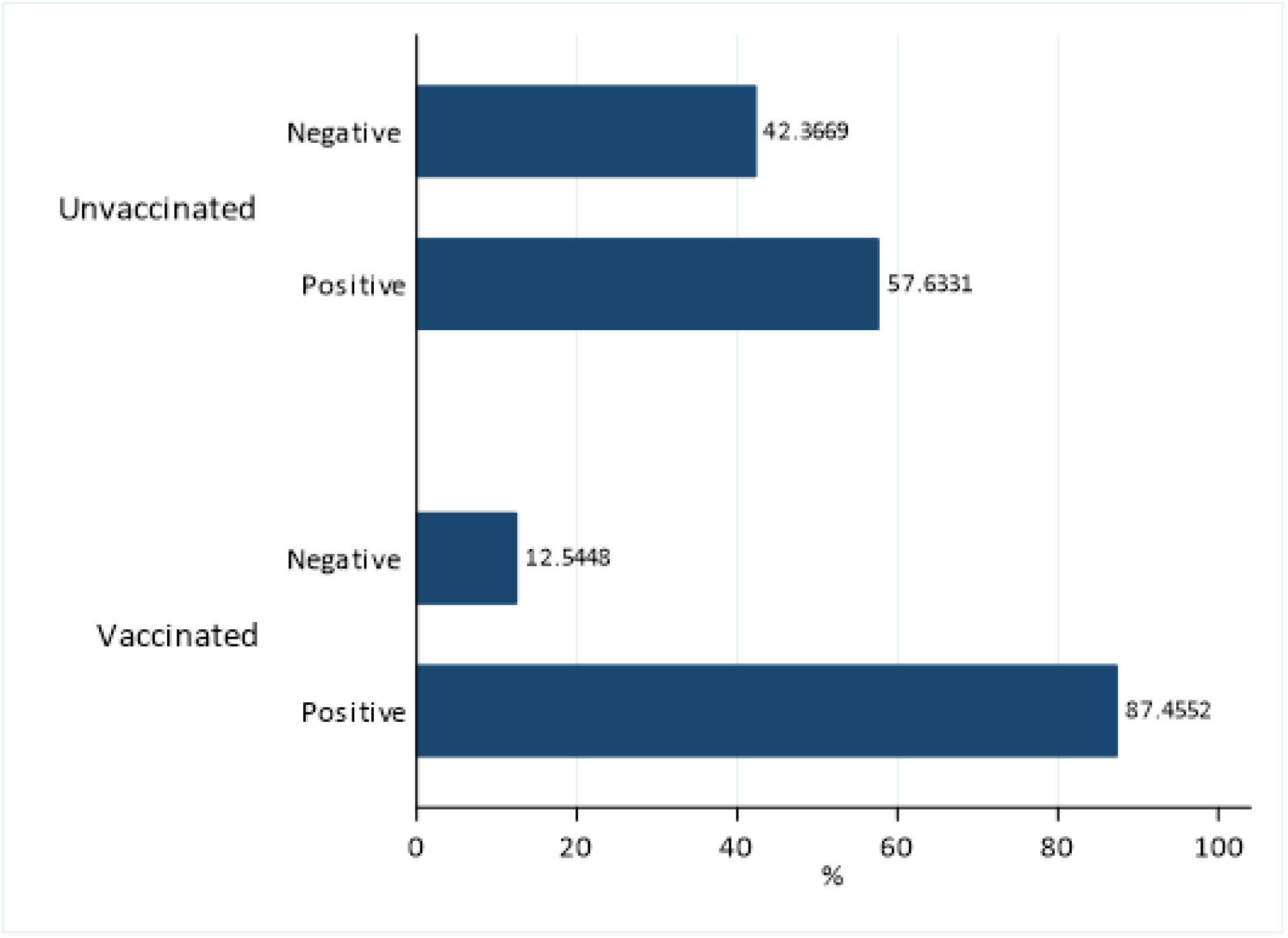
Graph showing the association between COVID-19 seropositivity (yes/no) and vaccination status.

**Supplementary Figure 2:**
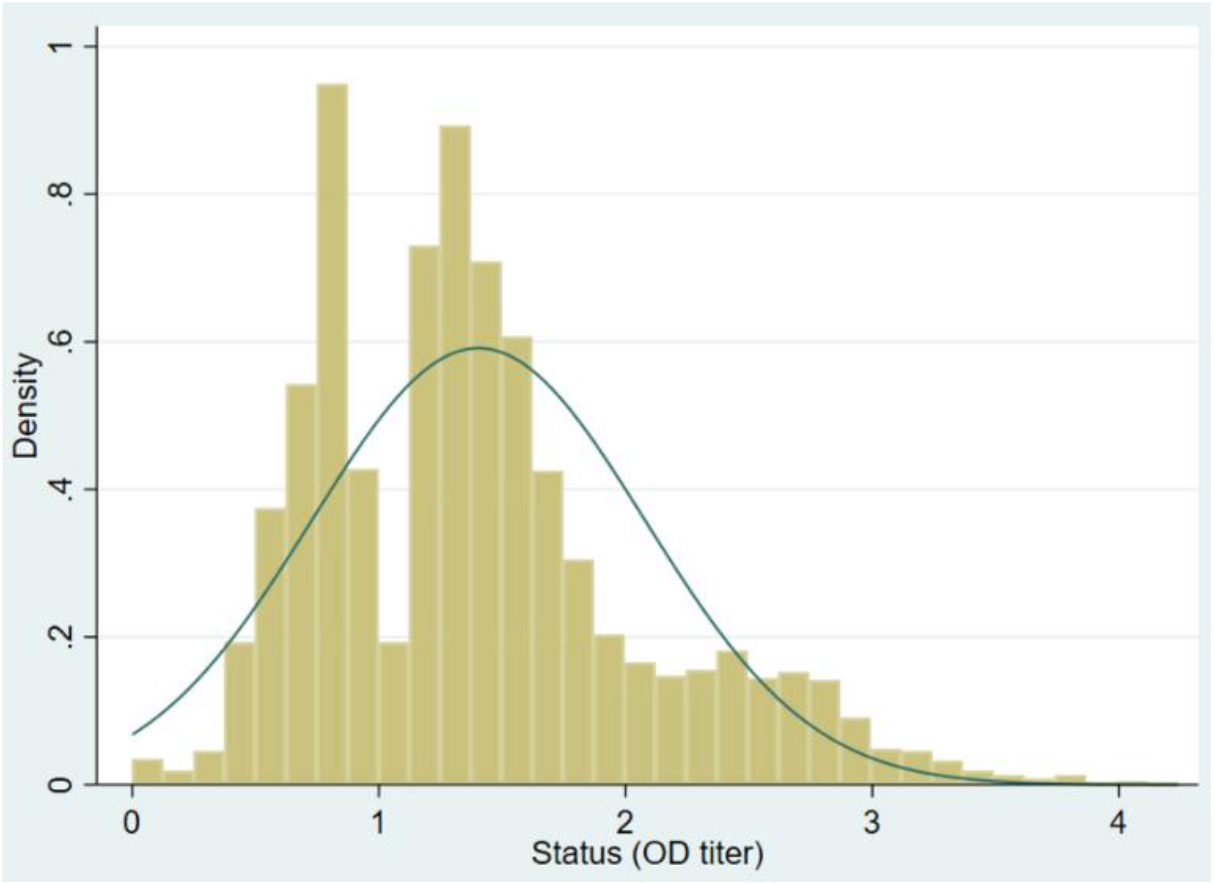
Graph showing the distribution of OD titer.

