## Supplemental Information for "Joint Impacts of Nutrition and Socioeconomic Status on SARS-CoV-2 Antibody Titer in a Prospective Birth Cohort in Brazil"

**Appendices**

**Supplementary Table 1.** Logistic regression.

|  | **Unvaccinated** | | **Vaccinated** | | **Combined** | |
| --- | --- | --- | --- | --- | --- | --- |
|  | **Unadjusted**  **OR (95% CI)** | **Adjusted#**  **OR (95% CI)** | **Unadjusted**  **OR (95% CI)** | **Adjusted#**  **OR (95% CI)** | **Unadjusted**  **OR (95% CI)** | **Adjusted#**  **OR (95% CI)** |
| **SES** |  |  |  |  |  |  |
| 1 (low) | Ref | Ref | Ref | Ref | Ref | Ref |
| 2 | .82 (.55, 1.24) | .75 (.46, 1.20) | 1.36 (.84, 2.18) | 1.24 (.74, 2.06) | 1.09 (.86, 1.38) | .95 (.67, 1.34) |
| 3 | .99 (.64, 1.51) | 1.12 (.69, 1.83) | 1.24 (.77, 1.99) | 1.33 (0.80, 2.22) | 1.06 (.83, 1.34) | 1.23 (.86, 1.76) |
| 4 | .92 (.60, 1.42) | .90 (.55, 1.47) | 1.55 (.94, 2.56) | 1.41 (.82, 2.41) | 1.08 (.85, 1.38) | 1.11 (.77, 1.59) |
| 5 (high) | 1.21 (.77, 1.89) | 1.11 (.66, 1.87) | 1.44 (.89, 2.35) | 1.49 (.88, 2.54) | 1.19 (.93, 1.52) | 1.27 (.88, 1.85) |
| **Stunting** |  |  |  |  |  |  |
| No | Ref | Ref | Ref | Ref | Ref | Ref |
| Yes | .75 (.50, 1.14) | .74 (.45, 1.20) | 1.08 (.67, 1.74) | 1.00 (.60, 1.68) | .94 (.74, 1.19) | .86 (.61, 1.21) |
| **Age (years)** | 2.80 (1.79, 4.37) | .87 (.48, 1.59) | 2.41 (1.49, 3.90) | .61 (.33, 1.12) | 2.85 (2.21, 3.67) | .74 (.48, 1.14) |
| **DTP (doses)** | 1.11 (1.01, 1.23) | 1.15 (1.03, 1.28) | 1.10 (.99, 1.23) | 1.07 (.95, 1.21) | 1.06 (1.00, 1.11) | 1.12 (1.03, 1.21) |
| **School entry (months)** | 1.33 (1.24, 1.42) | 1.41 (1.29, 1.55) | 1.50 (1.36, 1.65) | 1.60 (1.43, 1.80) | 1.61 (1.53, 1.70) | 1.48 (1.38, 1.60) |
| **Vaccine** |  |  |  |  |  |  |
| No | -- | -- | -- | -- | Ref | Ref |
| Yes | -- | -- | -- | -- | 5.12 (4.16, 6.32) | 3.72 (2.95, 4.71) |

 #Adjusted for all variables at the same time

**Supplementary Figure 1**: Graph showing the association between COVID-19 seropositivity (yes/no) and vaccination status.

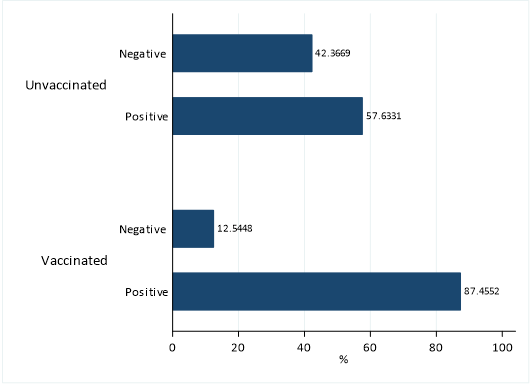

**Supplementary Figure 2:** Graph showing the distribution of OD titer.

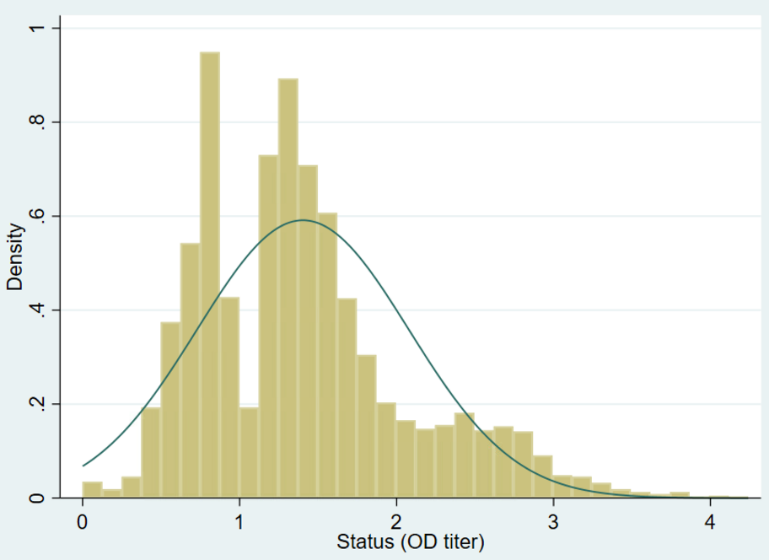
